# Completion of multi-dose COVID-19 vaccines and the associated factors among adolescents and adults in two urban informal settlements in Nairobi, Kenya

**DOI:** 10.1101/2024.11.04.24316733

**Authors:** Maurine Ng’oda, Jonathan Izudi, Collins Otieno, Daniel Mwanga, Richard E. Sanya, Abdhalah Ziraba

## Abstract

Completion of vaccine doses is essential for robust immunity and long-term protection against specific diseases. This study aimed to investigate factors associated with the completion of multi-dose COVID-19 vaccines (MDV) among adolescents and adults in two informal urban settlements in Nairobi, Kenya. We analyzed data from the Kenya Multisite Integrated Serosurveillance (KEMIS) project. We defined completion of MDV as receiving an additional COVID-19 vaccine dose between weeks 6 and 8 following the first COVID-19 vaccination shot. We applied the modified Poisson regression model with robust standard errors to determine the factors that were independently associated with the completion of MDV. We analyzed data from 402 individuals aged 14-90 years and found that the completion rate for MDV was 85.3%. In the adjusted analysis, participants aged ≥60 years (adjusted prevalence risk ratio [aPR] 1.30, 95% confidence interval [CI] 1.05-1.60) and those who recommended the COVID-19 vaccine to others (aPR 1.40, 95% CI 1.00-1.96) were significantly more likely to complete MDV. Individuals aged 25-59 years (aPR 1.22, 95% CI 0.99-1.50), those who perceived themselves as being at risk for COVID-19 (aPR 1.09, 95% CI 0.99-1.19), and those who had access to healthcare during the pandemic tended to have a higher completion of MDV. Overall, the MDV completion rate is relatively high, however, public health interventions should endeavor to target those being left behind such as younger individuals and those hesitant about vaccination.

## BACKGROUND

The COVID-19 pandemic posed unprecedented challenges to global public health, with vaccination emerging as one of the most effective tools to mitigate transmission and reduce severe outcomes such as hospitalization and mortality.^1^By August 12, 2024, 13.72 billion doses of COVID-19 vaccine had been administered globally. However, Africa had received the least number of doses (874.76 million), ^2^ revealing significant gaps in both vaccine access and uptake.^3,4^

In Kenya, the most commonly used COVID-19 vaccines included Oxford/AstraZeneca, Moderna, and Pfizer and each requires multiple doses to ensure complete immunization.^5^ The completion of multi-dose vaccine regimens is critical for eliciting stronger immunity and longer-term protection against severe COVID-19. The second dose significantly enhances the immune response, boosting the efficacy to over 90% in preventing severe disease.^6^ Failure to complete multi-dose regimens undermines this protection and potentially leads to vaccine failure, greater risks for infection, and reduced overall effectiveness of immunization programs including increased susceptibility to COVID-19 variants and reduced community-level immunity.^7^

Disparities in the completion of multi-dose vaccines for COVID-19 (MDV) exist, particularly in low-income and marginalized populations such as urban informal settlements.^8^ Urban informal settlements are often characterized by overcrowding, inadequate healthcare infrastructure, and limited access to essential services. In Kenya, an estimated 60% of the residents in Nairobi City reside in informal settlements. The overall COVID-19 vaccine uptake among this population is around 34% ^9^ but the completion of MDV is understudied. Understanding the completion of MDV is crucial for developing targeted interventions that improve COVID-19 vaccine uptake in such settings. We investigated the prevalence of completion of MDV and the associated factors among adolescents and adults in two large urban informal settlements in Nairobi, Kenya.

## MATERIALS AND METHODS

### Study setting

Data used in this paper comes from the Kenya Multisite Integrated Serosurveillance (KEMIS) for COVID-19 and Other Pathogens Project. The KEMIS Project was jointly implemented by the African Population and Health Research Center (APHRC), the Kenya Medical Research Institute Wellcome Trust Research Programme, and the Centers for Disease Control and Prevention (CDC). The project aimed to estimate the sero-prevalence of SARS-CoV-2 antibodies in the general population. It involved serosurveys conducted among residents of Kilifi, Nairobi, and Manyatta Health and Demographic Surveillance Systems (HDSS). The details of HDSS have been previously discussed.^9^ The analysis in this paper comes from the Nairobi site. The Nairobi Urban HDSS (NUHDSS) is established in Nairobi City County, and has consistently tracked a population of approximately 90,000 people each year spread across 30,000 households in two slum areas of Korogocho and Viwandani.^10^

### Study design and eligibility

Four cross-sectional sero-surveys were conducted between October 2020 and June 2024 on a random age-stratified sample of 850 NUHDSS residents totaling 3,400 observations. These samples consisted of 50 people from each 5-year age group between 15 and 64 years, 50 people aged 65 and above, and 100 children from each 5-year age group between 0 and 14 years. This resulted in 300 participants under 15 years old sufficient to estimate a 1% outcome measure rate with a 2% margin of error. For the 15-64 age group, 500 participants were deemed adequate to estimate an outcome rate of 3-5% within a 5% margin of error. The survey excluded individuals with bleeding disorders and other medical contraindications for venipuncture or capillary blood sample collection. Data was collected using structured questionnaires and laboratory analysis of blood samples for COVID-19 antibodies. The questionnaire captured sociodemographic information, information on likely exposure to SARS-Cov-2, and clinical information such as COVID-19 symptoms, childhood immunization, death of household members, and vaccination status. Vaccination status was verified using the vaccination certificate or text message confirmation from the national COVID-19 registry. Other information collected included access to health services during the pandemic and loss of income during COVID-19.

### Measurements and variables

The present study utilized data from the second and third survey rounds of the survey since the first round of the survey took place before the COVID-19 vaccine rollout in the country and the fourth round of the survey did not include key variables that were considered for the current analysis. We considered observations for individuals who were eligible for COVID-19 vaccination and who had received the first dose of MDV, namely AstraZeneca, Moderna, Pfizer, and Sinopharm. We excluded records for the following individuals: those ineligible for the COVID-19 vaccine based on age, who never received any COVID-19 vaccination, who never knew the vaccine they had received at first vaccination, and those who had missing vaccination information.

The primary outcome variable of interest was completion of MDV measured as a binary outcome and defined as the receipt of an additional dose of a COVID-19 vaccine between weeks 6 and 8 following the first dose of an MDV. The independent variables included age measured as the number of years lived and categorized as 14–24, 25–59, and ≥60 years to respectively denote adolescents and young people, middle-aged persons, and older persons; sex (male vs. female); religion measures as None, Muslim, Christians, and Others; education level was measured as none, primary, secondary, and post-secondary; exposure to a suspected COVID-19 case was measured as no, yes, probable, and not reported; access to healthcare during the COVID-19 pandemic was measured on a binary scale as yes or no; and perceived risk of COVID-19 infection was measured as no risk vs. at risk. Additionally, we assessed vaccine whether the participants were willing to recommend COVID-19 vaccination to others and whether they had any household member who had died from COVID-19.

### Statistical analysis

We summarized categorical variables as frequencies and proportions. Numerical variables were summarized using the mean and standard deviation as they were normally distributed. The Chi-square test was used to examine differences in MDV completion by the covariates at a 0.15 significance level in the bivariate analysis. We used the modified (robust) Poisson regression model to determine the factors independently associated with MDV completion adjusting for both clinically and socially relevant variables from the literature, statistically significant variables at the bivariate analysis, and robust standard errors as recommended by Cameron and Trivedi. We report both the unadjusted and adjusted prevalence risk ratios (PR) along with the 95% confidence intervals (CIs). Variables with p<0.05 were considered statistically significant in the multivariable modified Poisson regression analysis.

### Ethical considerations

The KEMIS project obtained ethical approval from the Kenya Medical Research Institute Scientific and Ethics Review Unit (Reference no. KEMRI/SERU/ CGMR-C/203/4085), The National Commission for Science, Technology and Innovation (License no. NACOSTI/P/21/13771), the Oxford Tropical Research Ethics Committee (Reference no. 44–20), and the London School of Hygiene and Tropical Medicine Research Ethics Committee (Reference no. 26950). Written parental/guardian consent was obtained for participants aged <18 years, accompanied by written assent for children aged 13 to 17 years.

## RESULTS

### Study profile

We considered data from 1,701 participants and excluded 1,295 that did not meet the eligibility criteria. Overall, we analyzed 402 participants of whom 343 (85.0%) completed MDV as shown in Figure 1.

**Figure 1.**
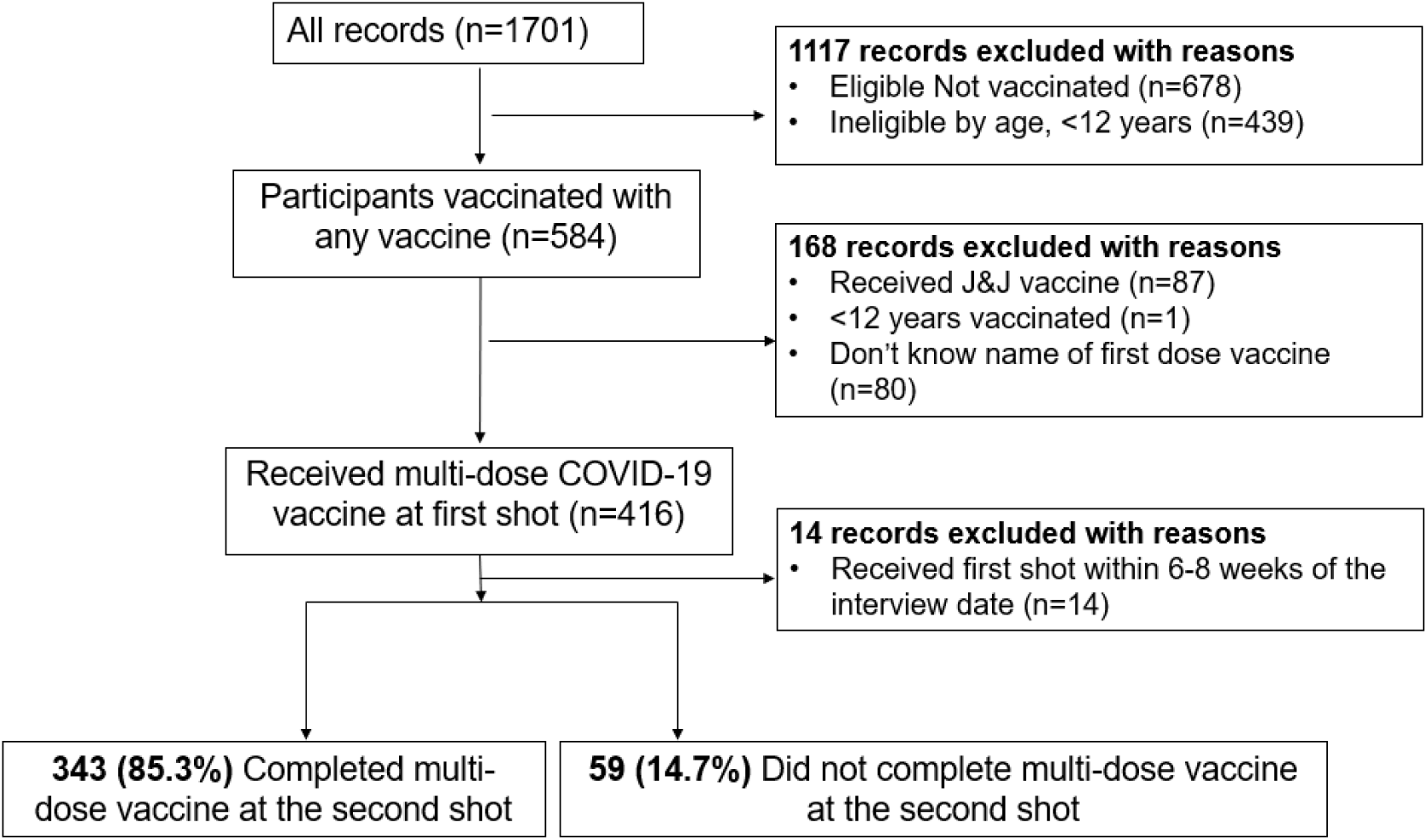
Study profile based on eligibility criteria.

### General characteristics of the participants

We present the general characteristics of the participants in Table 1. Of the 402 participants, 262 (65.2%) were from Viwandani study site, 264 (65.7%) were male, 254 (63.2%) were aged 25-59 years and the overall mean age was 48.5±15.7 332 (80.1%) participants had no known contact with confirmed/suspected COVID-19 cases, 260 (64.7%) perceived themselves as being at risk for infection, and 361 (89.8%) lacked access to healthcare during the pandemic.

**Table 1.**
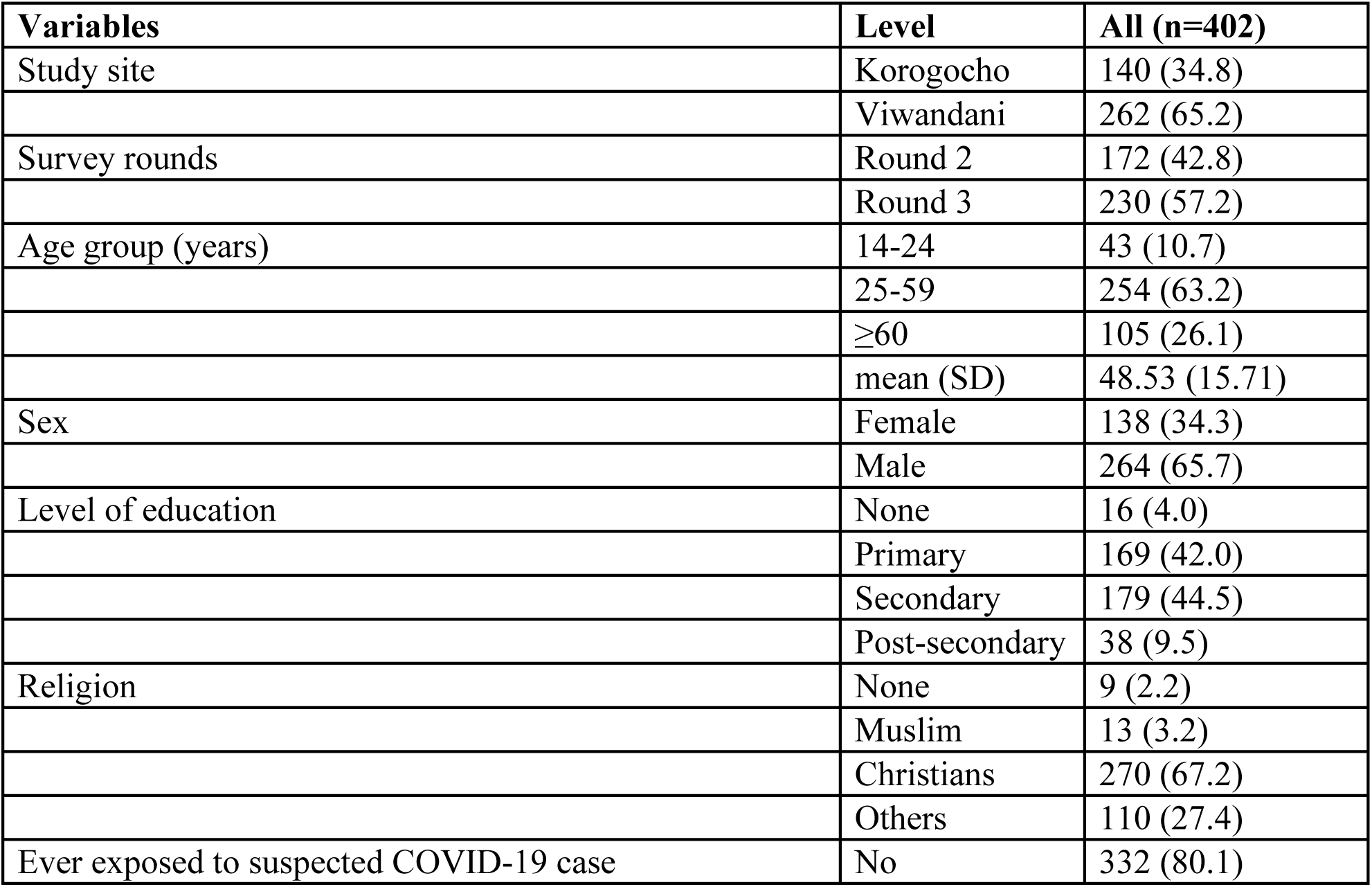
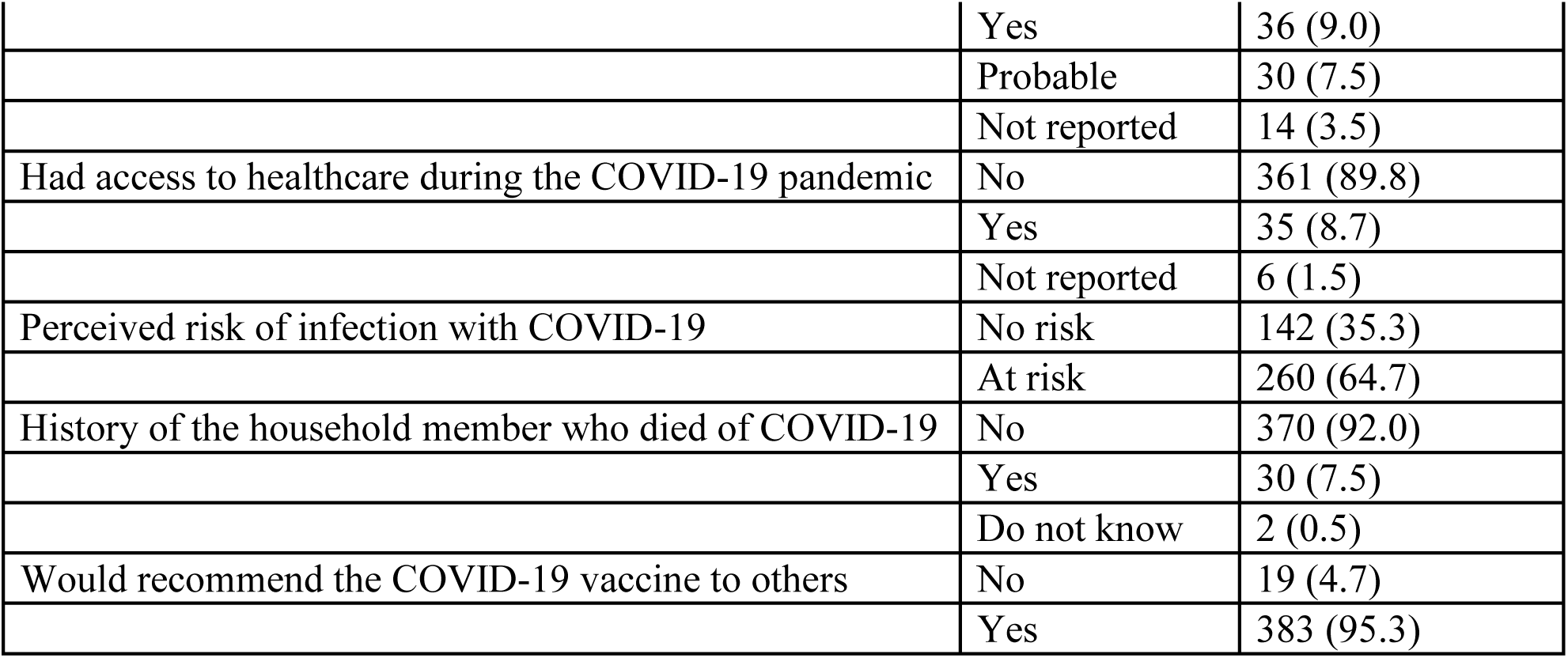
General characteristics of participants who received multi-dose COVID-19 vaccine.

| Variables | Level | All (n=402) |
| --- | --- | --- |
| Study site | Korogocho | 140 (34.8) |
|  | Viwandani | 262 (65.2) |
| Survey rounds | Round 2 | 172 (42.8) |
|  | Round 3 | 230 (57.2) |
| Age group (years) | 14-24 | 43 (10.7) |
|  | 25-59 | 254 (63.2) |
|  | ≥60 | 105 (26.1) |
|  | mean (SD) | 48.53 (15.71) |
| Sex | Female | 138 (34.3) |
|  | Male | 264 (65.7) |
| Level of education | None | 16 (4.0) |
|  | Primary | 169 (42.0) |
|  | Secondary | 179 (44.5) |
|  | Post-secondary | 38 (9.5) |
| Religion | None | 9 (2.2) |
|  | Muslim | 13 (3.2) |
|  | Christians | 270 (67.2) |
|  | Others | 110 (27.4) |
| Ever exposed to suspected COVID-19 case | No | 332 (80.1) |
|  | Yes | 36 (9.0) |
|  | Probable | 30 (7.5) |
|  | Not reported | 14 (3.5) |
| Had access to healthcare during the COVID-19 pandemic | No | 361 (89.8) |
|  | Yes | 35 (8.7) |
|  | Not reported | 6 (1.5) |
| Perceived risk of infection with COVID-19 | No risk | 142 (35.3) |
|  | At risk | 260 (64.7) |
| History of the household member who died of COVID-19 | No | 370 (92.0) |
|  | Yes | 30 (7.5) |
|  | Do not know | 2 (0.5) |
| Would recommend the COVID-19 vaccine to others | No | 19 (4.7) |
|  | Yes | 383 (95.3) |

### Type and timing of receipt of multi-dose COVID-19 vaccine (MDV)

Table 2 summarizes the types and timing of MDV at the first and second shots. At the first shot, 266 (66.2%) participants received the AstraZeneca vaccine, 61 (15.2%) received Pfizer, 74 (18.4%) received Moderna, and 1 (0.2%) received the Sinopharm vaccine. At the second shot, the number of participants that received the AstraZeneca vaccine dropped to 224 (55.7%), and that for Pfizer and Moderna dropped to 59 (14.7%) and 56 (13.9%) respectively. One participant received Sinopharm at the first and second shots. At the second shot, 59 (14.7%) participants did not return for their COVID-19 vaccine. Overall, 343 (85.3%) participants completed their MDV in this study.

**Table 2.** Type and timing of receipt of multi-dose COVID-19 vaccine (MDV)

| Variable | Level | Overall (n=402) |
| --- | --- | --- |
| COVID-19 vaccine at the first shot | AstraZeneca | 266 (66.2) |
|  | Pfizer | 61 (15.2) |
|  | Moderna | 74 (18.4) |
|  | Sinopharm | 1 (0.2) |
| COVID-19 vaccine at the second shot | AstraZeneca | 224 (55.7) |
|  | Pfizer | 59 (14.7) |
|  | J & J | 1 (0.2) |
|  | Moderna | 56 (13.9) |
|  | Sinopharm | 1 (0.2) |
|  | Don't know | 2 (0.5) |
|  | Never returned | 59 (14.7) |
| Completed multi-dose COVID-19 vaccine (MDV) | No | 59 (14.7) |
|  | Yes | 343 (85.3) |

### Completion of MDV by the socio-demographic characteristics and other factors

The completion of MDV varied across different sociodemographic characteristics and other influencing factors as shown in Table 3. Completion rates were high across both study sites, with 85.7% in Korogocho and 85.1% in Viwandani (p=0.871). Individuals aged ≥60 years had the highest completion rate at 91.4%, while those aged 14-24 had a lower completion rate at 69.8% (p=0.003). The mean age of completers was significantly higher (50.14 years) compared to non-completers (39.17 years, p<0.001). There were no significant differences in completion rates by sex, with males and females completing at rates of 85.6% and 84.8%, respectively (p=0.825). Educational level showed modest differences, with those having no education showing the highest completion rate (93.8%), though this was not statistically significant (p=0.504). Religion was not a significant factor, though Muslims reported a 100% completion rate compared to 86.7% among Christians (p=0.102). The perceived risk of COVID-19 infection was marginally associated with completion, as those perceiving themselves at risk had a higher completion rate (87.7%) compared to those perceiving no risk (81.0%, p=0.069). Finally, willingness to recommend the COVID-19 vaccine was significantly associated with completion; 86.4% of those who would recommend the vaccine completed MDV compared to 63.2% among those who would not recommend it (p=0.005).

**Table 3:**
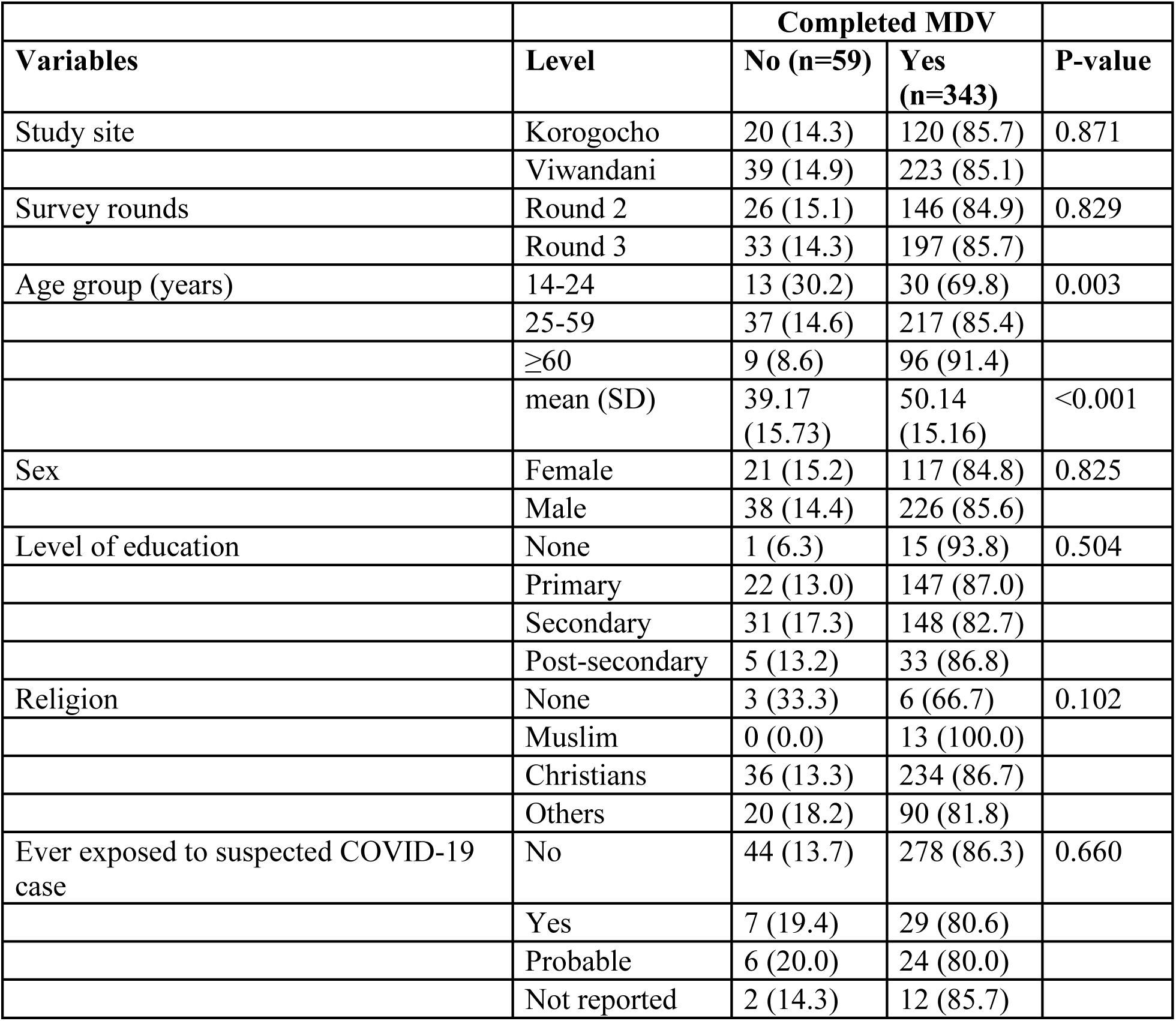
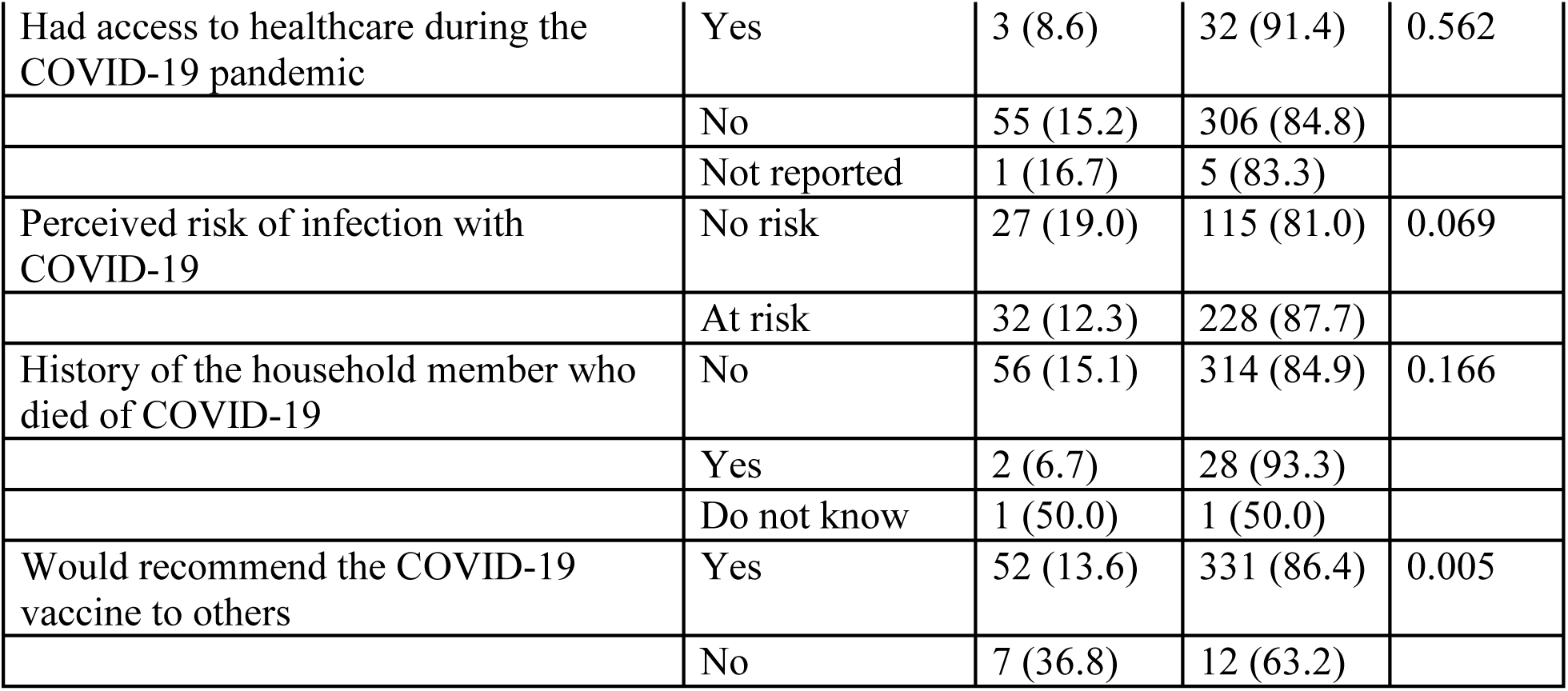
Completion of MDV by the socio-demographic characteristics and other factors.

| Variables | Level | Completed MDV |  | P-value |
| --- | --- | --- | --- | --- |
|  |  | No (n=59) | Yes (n=343) |  |
| Study site | Korogocho | 20 (14.3) | 120 (85.7) | 0.871 |
|  | Viwandani | 39 (14.9) | 223 (85.1) |  |
| Survey rounds | Round 2 | 26 (15.1) | 146 (84.9) | 0.829 |
|  | Round 3 | 33 (14.3) | 197 (85.7) |  |
| Age group (years) | 14-24 | 13 (30.2) | 30 (69.8) | 0.003 |
|  | 25-59 | 37 (14.6) | 217 (85.4) |  |
|  | ≥60 | 9 (8.6) | 96 (91.4) |  |
|  | mean (SD) | 39.17 (15.73) | 50.14 (15.16) | <0.001 |
| Sex | Female | 21 (15.2) | 117 (84.8) | 0.825 |
|  | Male | 38 (14.4) | 226 (85.6) |  |
| Level of education | None | 1 (6.3) | 15 (93.8) | 0.504 |
|  | Primary | 22 (13.0) | 147 (87.0) |  |
|  | Secondary | 31 (17.3) | 148 (82.7) |  |
|  | Post-secondary | 5 (13.2) | 33 (86.8) |  |
| Religion | None | 3 (33.3) | 6 (66.7) | 0.102 |
|  | Muslim | 0 (0.0) | 13 (100.0) |  |
|  | Christians | 36 (13.3) | 234 (86.7) |  |
|  | Others | 20 (18.2) | 90 (81.8) |  |
| Ever exposed to suspected COVID-19 case | No | 44 (13.7) | 278 (86.3) | 0.660 |
|  | Yes | 7 (19.4) | 29 (80.6) |  |
|  | Probable | 6 (20.0) | 24 (80.0) |  |
|  | Not reported | 2 (14.3) | 12 (85.7) |  |
| Had access to healthcare during the COVID-19 pandemic | Yes | 3 (8.6) | 32 (91.4) | 0.562 |
|  | No | 55 (15.2) | 306 (84.8) |  |
|  | Not reported | 1 (16.7) | 5 (83.3) |  |
| Perceived risk of infection with COVID-19 | No risk | 27 (19.0) | 115 (81.0) | 0.069 |
|  | At risk | 32 (12.3) | 228 (87.7) |  |
| History of the household member who died of COVID-19 | No | 56 (15.1) | 314 (84.9) | 0.166 |
|  | Yes | 2 (6.7) | 28 (93.3) |  |
|  | Do not know | 1 (50.0) | 1 (50.0) |  |
| Would recommend the COVID-19 vaccine to others | Yes | 52 (13.6) | 331 (86.4) | 0.005 |
|  | No | 7 (36.8) | 12 (63.2) |  |

**Table 4.**
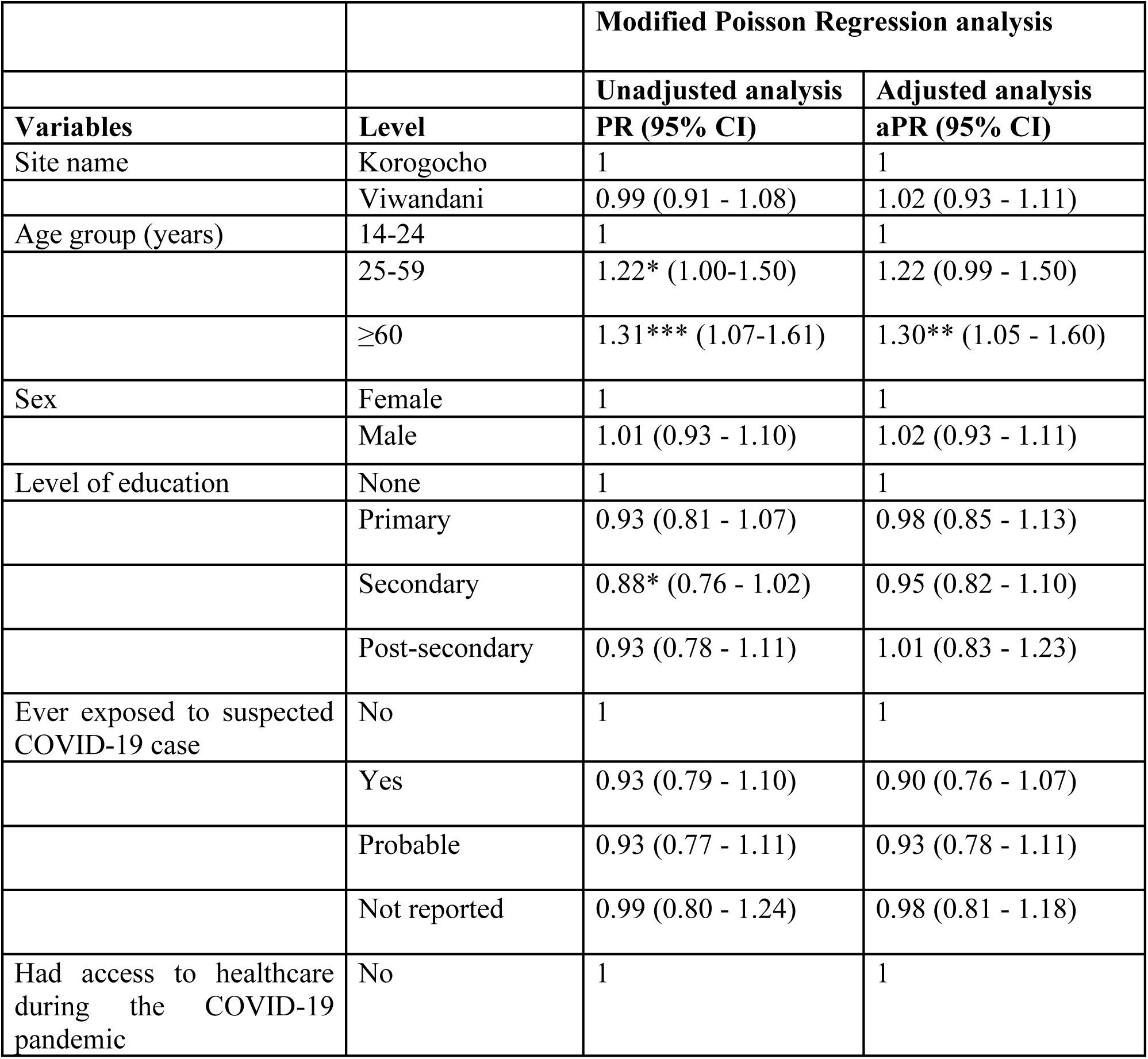
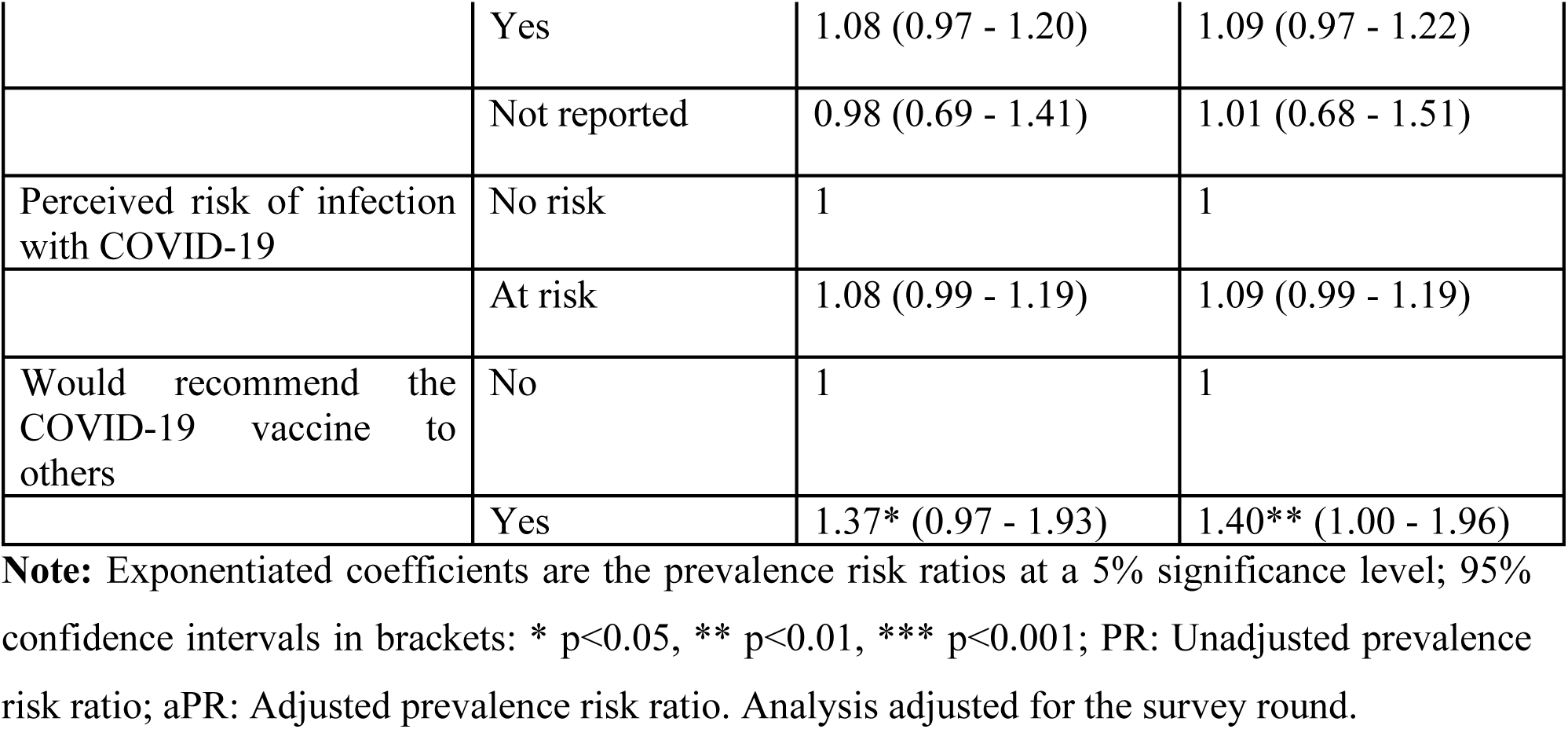
Factors associated with the completion of MDV.

|  |  | Modified Poisson Regression analysis |  |
| --- | --- | --- | --- |
|  |  | Unadjusted analysis | Adjusted analysis |
| Variables | Level | PR (95% CI) | aPR (95% CI) |
| Site name | Korogocho | 1 | 1 |
|  | Viwandani | 0.99 (0.91 - 1.08) | 1.02 (0.93 - 1.11) |
| Age group (years) | 14-24 | 1 | 1 |
|  | 25-59 | 1.22* (1.00-1.50) | 1.22 (0.99 - 1.50) |
|  | ≥60 | 1.31*** (1.07-1.61) | 1.30** (1.05 - 1.60) |
| Sex | Female | 1 | 1 |
|  | Male | 1.01 (0.93 - 1.10) | 1.02 (0.93 - 1.11) |
| Level of education | None | 1 | 1 |
|  | Primary | 0.93 (0.81 - 1.07) | 0.98 (0.85 - 1.13) |
|  | Secondary | 0.88* (0.76 - 1.02) | 0.95 (0.82 - 1.10) |
|  | Post-secondary | 0.93 (0.78 - 1.11) | 1.01 (0.83 - 1.23) |
| Ever exposed to suspected COVID-19 case | No | 1 | 1 |
|  | Yes | 0.93 (0.79 - 1.10) | 0.90 (0.76 - 1.07) |
|  | Probable | 0.93 (0.77 - 1.11) | 0.93 (0.78 - 1.11) |
|  | Not reported | 0.99 (0.80 - 1.24) | 0.98 (0.81 - 1.18) |
| Had access to healthcare during the COVID-19 pandemic | No | 1 | 1 |
|  | Yes | 1.08 (0.97 - 1.20) | 1.09 (0.97 - 1.22) |
|  | Not reported | 0.98 (0.69 - 1.41) | 1.01 (0.68 - 1.51) |
| Perceived risk of infection with COVID-19 | No risk | 1 | 1 |
|  | At risk | 1.08 (0.99 - 1.19) | 1.09 (0.99 - 1.19) |
| Would recommend the COVID-19 vaccine to others | No | 1 | 1 |
|  | Yes | 1.37* (0.97 - 1.93) | 1.40** (1.00 - 1.96) |

**Note:** Exponentiated coefficients are the prevalence risk ratios at a 5% significance level; 95% confidence intervals in brackets: \* p<0.05, \*\* p<0.01, \*\*\* p<0.001; PR: Unadjusted prevalence risk ratio; aPR: Adjusted prevalence risk ratio. Analysis adjusted for the survey round.

### Factors associated with the completion of MDV at the unadjusted and adjusted analyses

In the unadjusted analysis, participants aged 25-59 years (PR 1.22, 95% CI 1.00-1.50) and ≥60 years (PR 1.31, 95% CI 1.07-1.61) were more likely to complete MDV compared to those aged 14-24 years. Being a male compared to a female was not associated with completion of MDV (PR 1.01, 95% CI 0.93-1.10). Education levels, namely primary (PR 0.93, 95% CI 0.81-1.07), secondary (PR 0.88, 95% CI 0.76-1.02), and post-secondary education (PR 0.93, 95% CI 0.78-1.11) showed no significant association with completion of MDV compared to no formal education. Participants who perceived themselves as being at risk for COVID-19 (PR 1.08, 95% CI 0.99-1.19) and those who would recommend the vaccine to others (PR 1.37, 95% CI 0.97-1.93) tended to have a higher likelihood of completion of MDV.

In the adjusted analysis, participants aged ≥60 years (aPR 1.30, 95% CI 1.05-1.60) and those who would recommend the COVID-19 vaccine to others (aPR 1.40, 95% CI 1.00-1.96) were more likely to complete MDV. We found a borderline statistically significant association between being aged 25-59 years compared to 14-24 years (aPR 1.22, 95% CI 0.99 - 1.50), access to healthcare during the COVID-19 pandemic (aPR 1.09, 95% CI 0.97 - 1.22), and perceived risk of COVID-19 (aPR 1.09, 95% CI 0.99 −1.19).

## DISCUSSION

We assessed the factors associated with the completion of MDV in Nairobi’s informal settlements. Our findings indicate that 8 in every 10 individuals in the informal settlements who ever received a COVID-19 vaccine completed their MDV. Completion of MDV is more likely for middle-aged (25-59 years) and older adults (60+ years) individuals compared to younger participants (14-24 years) and among those reporting willingness to recommend COVID-19 vaccination to others. Furthermore, individuals who perceived themselves at risk for COVID-19 and those who had access to health services during the pandemic tended to have a higher likelihood of completing their MDV.

Age has been shown in previous studies to be a significant determinant of vaccine uptake ^11,12^ and our findings align with this evidence. The high completion rate of MDV among middle-aged individuals, who comprise the majority of the working population, may have been likely influenced by the COVID-19 restrictions that largely affected city employees. Across urban areas, including Nairobi, several employers mandated COVID-19 vaccination as a requirement for continued employment, especially in sectors involving close contact with others, such as hospitality, transport, and retail.^13,14^ These mandates might have incentivized middle-aged workers to complete their MDVregimens in order to retain their jobs and avoid financial instability Compared to younger persons, middle-aged and older individuals might have prioritized vaccination in order to reduce the risk of transmission of SARS-COV-2 (the virus causing COVID-19) at the household level. Diesel and colleagues (2021) argued that older adults are more likely to complete their COVID-19 vaccine regimens due to a greater awareness of the health risks associated with COVID-19.^15^ One study indicated that older adults prioritize vaccination due to their higher perceived risk of severe outcomes associated with COVID-19.^16^

We found that participants who would recommend the COVID-19 vaccine to others are more likely to complete their MDV compared to those who would not recommend the COVID-19 vaccine to others. This finding highlights the critical role of vaccine confidence in shaping health behavior. Vaccine-non-hesitant individuals often have a higher level of trust in the safety and efficacy of vaccines, which motivates them to get vaccinated and advocate for others to equally get vaccinated.^17^ This positive attitude toward vaccination likely translates into greater adherence to completing the full vaccine schedule since such individuals likely appreciate the benefits of vaccination. Contrastingly, vaccine-hesitant individuals may harbour doubts about vaccine safety or question its necessity and hence are less likely to commit to completing the multi-dose regimens.^17^ This finding emphasizes the importance of addressing vaccine hesitancy through public health messaging and community engagement, particularly in settings like informal urban settlements where misinformation and distrust are prevalent.

Individuals with a perceived risk of COVID-19 infection showed a marginally higher tendency to complete their MDV. We argue that feeling at risk of COVID-19 infection, by itself, may not sufficiently motivate an individual to adhere to vaccination schedules but rather other factors such as the perceived benefits of full vaccination, easy access to vaccines, trust in the health system, and influences from the social networks.^18–20^ Even though some individuals might acknowledge that they are at risk for COVID-19, they may still hesitate to get vaccinated due to concerns about vaccine side effects, doubts about its effectiveness, or general mistrust of health interventions.^20,21^ Moreover, the lack of significant findings might suggest that people often need more than just a sense of risk to act. Public health theories such as the Health Belief Model highlight that while perceived risk can increase awareness, it often takes “cues to action” to trigger a response.^22^ These cues could include reminders from health professionals, public health campaigns, or healthy public policies such as vaccine mandates.

There are also other important dynamics at play such as vaccine fatigue.^23^ Some people might have been eager to get their initial dose but then become reluctant to receive subsequent doses due to misinformation or concerns after the first vaccination. This highlights that even when perceived risk is high, other competing factors may prevent people from completing their vaccination schedule.

Structural barriers such as limited access to vaccines, financial constraints, or the inability to take time off work also negatively affect people in urban slum environments regarding vaccination uptake. In urban informal settlements, basic needs such as food or income are prioritized over long-term health needs such as vaccination. Overall, public health strategies that focus exclusively on raising awareness of the risks associated with COVID-19 may not be effective in improving vaccine completion.

This study has numerous strengths and some limitations to note. The use of a modified Poisson regression analysis prevented the overestimation of associations between the independent variables and the outcome, hence providing better estimates. The study was conducted in only two informal settlements in Nairobi so the findings may not generalize to other urban slums. Missing data, particularly from participants who were unaware of the type of vaccine they had received at the first shot might have led to either under or overestimation of the completion of MDV. Despite these limitations, the study provides credible findings about the completion of MDV in urban informal settlements where access to healthcare remains problematic and individuals are at a higher risk for diseases, complications, and mortality.

We conclude that the completion rate for MDV is relatively high. However, to further enhance vaccination coverage, public health interventions should focus on targeted education, emphasizing the importance of completing the full vaccination schedule. These efforts should prioritize younger individuals and those displaying vaccine hesitancy, ensuring that critical gaps in coverage are addressed and sustained immunity is achieved within the population.

## ACKNOWLEDGEMENTS

This work is part of the African Population and Health Research Center’s commitment to advancing public health research in Africa, particularly in understanding and addressing health challenges within underserved communities. We are deeply grateful to our collaborators, the KEMRI Wellcome Trust Research Program and the Africa CDC, for their invaluable partnership in the KEMIS project. We also wish to thank the dedicated field data collectors and the community members who participated in the study, whose contributions were essential to this work. APHRC had no role in the study’s conceptualization, design, conduct, data analysis, or the decision to submit and publish the manuscript. The contents are solely the responsibility of the authors.

## AUTHOR DETAILS

Maurine Ng’oda; African Population and Health Research Center, Nairobi, Kenya.

Jonathan Izudi; African.

Collins Otieno; African.

Daniel Mwanga; African.

Richard E. Sanya; African Population and Health Research Center, Nairobi, Kenya.

Abdhalah Ziraba; African Population and Health Research Center, Nairobi, Kenya.

## DATA AVAILABILITY

All the data used in this manuscript are archived at https://microdataportal.aphrc.org/index.php/catalog/central and can be formally requested for through the institutional guidelines.

## AUTHOR DECLARATION

All authors have seen and approved this manuscript.

## REFERENCES

1. Kim D, Lee YJ. Vaccination strategies and transmission of COVID-19: Evidence across advanced countries. J Health Econ. 2022;82(January):102589. doi:10.1016/j.jhealeco.2022.102589

2. Our World in Data. Total COVID-19 vaccine doses administered.

3. Doshi RH, Nsasiirwe S, Dahlke M, et al. COVID-19 Vaccination Coverage — World Health Organization African Region, 2021–2023. MMWR Morb Mortal Wkly Rep. 2024;73(14):307–311. doi:10.15585/mmwr.mm7314a3

4. Jacobs C, Musonda NC, Tembo D, et al. Gender disparities and associated factors to intention to getting a second dose of COVID-19 AstraZeneca vaccine among adult populations in selected facilities of Lusaka, Zambia. PLOS Glob Public Heal. 2022;2(7):e0000265. doi:10.1371/journal.pgph.0000265

5. Solante R, Alvarez-Moreno C, Burhan E, et al. Expert review of global real-world data on COVID-19 vaccine booster effectiveness and safety during the omicron-dominant phase of the pandemic. Expert Rev Vaccines. 2023;22(1):1–16. doi:10.1080/14760584.2023.2143347

6. García-Montero C, Fraile-Martínez O, Bravo C, et al. An updated review of sars-cov-2 vaccines and the importance of effective vaccination programs in pandemic times. Vaccines. 2021;9(5):1–22. doi:10.3390/vaccines9050433

7. Crutcher M, Seidler PM. Maximizing completion of the two-dose COVID-19 vaccine series with aid from infographics. Vaccines. 2021;9(11). doi:10.3390/vaccines9111229

8. Ekezie W, Awwad S, Krauchenberg A, et al. Access to Vaccination among Disadvantaged, Isolated and Difficult-to-Reach Communities in the WHO European Region: A Systematic Review. Vaccines. 2022;10(7). doi:10.3390/vaccines10071038

9. Kagucia EW, Ziraba AK, Nyagwange J, et al. SARS-CoV-2 seroprevalence and implications for population immunity: Evidence from two Health and Demographic Surveillance System sites in Kenya, February–December 2022. Influenza Other Respi Viruses. 2023;17(9):1–11. doi:10.1111/irv.13173

10. Beguy D, Elung’ata P, Mberu B, et al. Health & Demographic Surveillance System Profile: The Nairobi Urban Health and Demographic Surveillance System (NUHDSS). Int J Epidemiol. 2015;44(2):462–471. doi:10.1093/ije/dyu251

11. Amoah JO, Abraham SA, Adongo CA, et al. Determinants of COVID-19 vaccine uptake: evidence from a vulnerable global South setting. BMC Res Notes. 2024;17(1):1–7. doi:10.1186/s13104-024-06736-5

12. Pijpers J, van Roon A, van Roekel C, et al. Determinants of COVID-19 Vaccine Uptake in The Netherlands: A Nationwide Registry-Based Study. Vaccines. 2023;11(9):1409. doi:10.3390/VACCINES11091409/S1

13. Lines K, Sebbanja JA, Dzimadzi S, et al. Covid-19 Vaccine Rollout: Challenges and Insights from Informal Settlements. 2022;53(3). doi:10.1177/09562478221149876

14. Rothstein MA, Parmet WE, Reiss DR. Employer-Mandated Vaccination for COVID-19. Am J Public Health. 2021;111(6):1061–1064. doi:10.2105/AJPH.2020.306166

15. Diesel J, Sterrett N, Dasgupta S, et al. COVID-19 Vaccination Coverage Among Adults — United States, December 14, 2020–May 22, 2021. Morb Mortal Wkly Rep. 2021;70(25):922–927. doi:10.15585/mmwr.mm7025e1

16. Bhanu C, Gopal DP, Walters K, Chaudhry UAR. Vaccination uptake amongst older adults from minority ethnic backgrounds: A systematic review. PLoS Med. 2021;18(11):1–17. doi:10.1371/journal.pmed.1003826

17. Naranjo D, Kimball E, Nelson J, et al. Differences in perceptions and acceptance of COVID-19 vaccination between vaccine hesitant and non-hesitant persons. PLoS One. 2023;18(9 September):1–15. doi:10.1371/journal.pone.0290540

18. Lin Y, Hu Z, Zhao Q, Alias H, Danaee M, Wong LP. Understanding COVID-19 vaccine demand and hesitancy: A nationwide online survey in China. PLoS Negl Trop Dis. 2020;14(12):e0008961. doi:10.1371/journal.pntd.0008961

19. Roberts-McCarthy E, Buck PO, Smith-Ray RL, et al. Factors associated with receipt of mRNA-1273 vaccine at a United States national retail pharmacy during the COVID-19 pandemic. Vaccines. 2023;41(29):4257–4266. doi:10.1016/j.vaccine.2023.03.076

20. Osur J, Muinga E, Carter J, Kuria S, Hussein S, Ireri EM. COVID-19 vaccine hesitancy: Vaccination intention and attitudes of community health volunteers in Kenya. PLOS Glob Public Heal. 2022;2(3 March):1–19. doi:10.1371/journal.pgph.0000233

21. Biswas MR, Alzubaidi MS, Shah U, Abd-Alrazaq AA, Shah Z. A scoping review to find out worldwide covid-19 vaccine hesitancy and its underlying determinants. Vaccines. 2021;9(11):1–20. doi:10.3390/vaccines9111243

22. Rukchart N, Hnuploy K, Eltaybani S, et al. Prevalence and determinants of COVID-19 vaccine acceptance among vulnerable populations in Thailand: An application of the health belief model. Heliyon. 2024;10(4):e26043. doi:10.1016/j.heliyon.2024.e26043

23. Stamm TA, Partheymüller J, Mosor E, et al. Determinants of COVID-19 vaccine fatigue. Nat Med. 2023;29(5):1164–1171. doi:10.1038/s41591-023-02282-y

